# Classifying chronic kidney disease from non-invasive questionnaire features in adults with long-standing diabetes in Bangladesh: model development, internal validation and the limits of small-sample evidence

**DOI:** 10.64898/2026.11.19.26368470

**Authors:** Ahmed Anan, Umma Tansina Arshi, Rafi Nazrul Islam

## Abstract

**Background:** Chronic kidney disease (CKD) is a common complication of diabetes, and in Bangladesh laboratory testing for it is not always available. Questionnaire answers could help decide whom to test first, but small machine-learning studies often report optimistic accuracy.

**Methods:** We analysed a public dataset of 400 adults with at least 10 years of diabetes attending a tertiary hospital in Dhaka, 185 with a hospital diagnosis of CKD. A pre-specified ridge logistic regression using 20 non-invasive features was developed and internally validated with nested cross-validation (five folds, ten repeats), compared with lasso, random forest and gradient boosting, and assessed for discrimination, calibration and net benefit. Simulations examined case-enriched sampling and single train/test splits.

**Results:** The area under the receiver operating characteristic curve (AUC) was 0.75 (95% confidence interval 0.70–0.80; optimism-corrected 0.76). Flexible algorithms were not detectably better (differences −0.005 to +0.006). Calibration within the sample was good (slope 1.06, 0.79–1.33), and testing the highest-scoring 30% of patients would have found 48% of CKD cases. The sample was smaller than the 883 patients recommended. Urinary infection, body-mass index, insulin use, diabetes duration and pain-killer use were positively associated with CKD and diet adherence negatively; regular walking and hypertension showed directions opposite to clinical expectation, consistent with features recorded after diagnosis. The yearly history added no reliable information. In simulation, case-enriched sampling preserved the AUC but overstated absolute risk (mean predicted risk 0.37 at a true prevalence of 0.10). Over 300 random 80/20 splits of the same data, the test AUC ranged from 0.61 to 0.87; reporting the best of ten splits would overstate it by about 0.07.

**Conclusions:** Questionnaire features moderately distinguished patients with an existing CKD diagnosis. Whether they identify patients before diagnosis requires external validation in patients assessed before CKD status is known. The study was not registered.

## Introduction

Diabetes affected an estimated 13.9 million adults in Bangladesh in 2024, 13.2% of the adult population, and about two in five of them were undiagnosed [1]. Diabetes is the leading cause of chronic kidney disease (CKD) worldwide [2], CKD is common in South Asia [3], and reduced kidney function is frequent among patients with type 2 diabetes attending a tertiary diabetes hospital in Dhaka [4]. Guidelines recommend at least annual testing of serum creatinine and urinary albumin in people with diabetes [5], but in low-resource settings these tests are not always available or affordable, and many patients are tested late. A tool that ranks patients by their likelihood of CKD from information that can be collected in a conversation could help clinics decide whom to test first. Simple questionnaire-based scores have been developed for this purpose in other populations [6].

Machine-learning studies of CKD often report very high accuracy, but they are typically based on small datasets of a few hundred patients, such as the widely used public dataset of 400 records [7], and are rarely validated in clinical settings or in diverse populations [8]. Across clinical prediction research, studies using machine learning are frequently at high risk of bias [9], flexible algorithms seldom outperform logistic regression [10], and leakage between training and test data inflates reported performance [11]. Small samples add a further problem: estimates from a single train/test split carry wide error bars [12], models developed on too few patients are unstable [13], and minimum sample sizes depend on the number of candidate predictors, the outcome prevalence and the anticipated model fit rather than on a fixed number of events per variable [14,15].

A public dataset collected at BIRDEM General Hospital in Dhaka records 20 non-invasive features, covering demographics, diabetes history and treatment, comorbidities, urinary symptoms, medication and lifestyle, for 400 adults with long-standing diabetes, 185 of whom had CKD [16,17]. These features can be collected without laboratory tests, but the dataset also has properties that shape what any model trained on it can show: it is small, it contains far more patients with CKD than a clinic population would, and the patients were interviewed once, after many of them had been diagnosed.

The aims of this study were therefore threefold. First, to develop and internally validate a pre-specified penalised logistic regression model that classifies CKD status from these 20 features, to compare it with more flexible algorithms, and to quantify its discrimination, calibration and, as an illustration, how it would rank patients for laboratory testing, with their uncertainty. Second, to examine which features carry information and whether their associations agree with clinical knowledge. Third, to quantify two methodological issues that affect small clinical datasets in general: how case-enriched sampling changes predicted probabilities, and how much a single train/test split can mislead at this sample size. Two secondary questions asked whether the pattern of diabetes management and the ten-year yearly history add information beyond the current profile.

## Materials and methods

### Study design and reporting

This is a secondary analysis of a published, de-identified dataset. We developed and internally validated a model that classifies CKD status in adults with long-standing diabetes from 20 non-invasive features, and we used the same data to examine how the sampling design and the evaluation procedure affect the evidence such models can provide. Reporting follows the TRIPOD+AI statement [18]; the completed checklist is provided in S1 Checklist.

The analysis plan, including the primary model, the primary metric and all secondary questions, was written into the analysis code (protocol version 1.0) and fixed before the final analysis was run; any later change would have been recorded in a deviation log, which remained empty. Exploratory analyses of the same dataset had been carried out before the plan was fixed, so the plan is not a prospectively registered protocol. The study was not registered, and the protocol file is available with the code. Patients and the public were not involved in the design, conduct or reporting of this analysis.

### Data source and participants

The dataset was collected at BIRDEM General Hospital, a tertiary diabetes hospital in Dhaka, Bangladesh, and is described in detail elsewhere [16,17]. In brief, adults who had lived with diabetes for at least 10 years and held a complete diabetes record book were interviewed with a structured questionnaire during routine hospital visits between March and June 2024. The questionnaire was developed with a supervising nephrologist and piloted twice before use, and answers were cross-checked against each patient’s record book. Each patient contributes 10 consecutive yearly records, and the last year of each record is the collection year. We analysed version 2 of the public dataset (SHA-256 fingerprint 9cef0eb4…3618, verified at every run). Before analysis, the code checked the structural properties that the analysis relies on: 400 patients with exactly 10 records each, no missing values, diabetes duration and age increasing by one year per record, and a CKD indicator that never reverts from 1 to 0.

### Outcome

The outcome was a diagnosis of CKD at any point in the 10-year record, as documented in each patient’s hospital-issued diabetes record book, in which physicians record CKD status on the basis of standard laboratory tests at regular check-ups. The laboratory values themselves are not part of the dataset, so CKD stage could not be analysed [16]. The dataset codes the diagnosis as a yearly indicator that changes from 0 to 1 in the year of diagnosis and remains 1 afterwards; no patient had CKD in the first year of the record.

### Predictors

The 20 candidate predictors were: gender, job exertion level (normal, intermediate, heavy), family history of diabetes, diabetes duration, age, body-mass index, adherence to the suggested diet, oral diabetes medicine, insulin, hypertension, heart disease, sufficient sleep (7–9 h per night), sufficient water intake (more than 2 L per day), smoking, zarda or betel leaf use, regular walking (at least 30 min per day), urinating properly, urinary infection or other urological issue, regular pain-killer use, and daily calorie intake (definitions and coding in Table A in S1 Appendix). Hypertension was recorded as present when the record book showed persistently high blood pressure (at least 140/90 mmHg) treated with antihypertensive medication; the drug class was not recorded. Calorie intake was estimated from the reported typical diet using standard dietary portion values [16]. Height, weight, average weight and average age were not used, because body-mass index is derived from height and weight and the averaged columns summarise the whole record. All 20 features were collected in the same interview and were treated alike in the analysis. Because CKD status was documented in the hospital records that were consulted during the interview, predictor assessment was not blinded to the outcome. Predictor values were taken from the first year of the record, when every patient was free of a CKD diagnosis. This choice does not make the analysis prospective. Several features are recorded as one value for the whole record (gender, family history, body-mass index, which is averaged over the 10 years, and calorie intake), urinary infection is a history over the 10-year period, job exertion describes the working life, and hypertension reflects blood pressure over the record. Most of the remaining features rarely change from year to year. The predictor values therefore largely describe patients as recorded at the collection interview, which for most patients with CKD took place after the diagnosis. We accordingly treat the task as classification of current CKD status and interpret associations as cross-sectional.

Binary features were coded 1 (present) and 0 (absent), job exertion as an ordinal score from 1 to 3, and continuous features on their original scales; all features were standardised inside the modelling pipeline using the training data only. There were no missing values, because complete records were an inclusion criterion.

### Models

The primary model, pre-specified because it is a regression model suited to small samples, was ridge-penalised (L2) logistic regression [19]. Its regularisation strength (inverse penalty C in {0.01, 0.03, 0.1, 0.3, 1, 3}) was chosen by 3-fold cross-validation on the training data, minimising log-loss so that tuning rewards well-calibrated probabilities as well as ranking. Comparators were lasso (L1) logistic regression [20] (C in {0.03, 0.1, 0.3, 1, 3}), a random forest [21] (200 trees; minimum leaf size 3 or 10; 50% of features or the square root of the number of features per split) and histogram-based gradient boosting [22] (150 iterations, L2 regularisation 1.0, minimum leaf size 15; maximum depth 2 or 3; learning rate 0.03 or 0.08). Two baselines bounded the comparison: a prevalence-only model, which assigns every patient the training prevalence, and a ridge model with age and diabetes duration only. Search grids were kept small on purpose, because large searches mainly fit noise at this sample size. No model was selected on the evaluation data; the comparators were evaluated to show whether more flexible algorithms changed the conclusions. No resampling or reweighting for class imbalance was used, because the two classes were of similar size. The model output is a probability; no classification threshold was chosen, and tertiles of the predicted probability were used only to describe observed CKD proportions.

### Internal validation

Performance was estimated by nested cross-validation [23]. The outer loop was stratified 5-fold cross-validation repeated 10 times (50 test folds of 80 patients, each with 320 training patients); the inner loop was the 3-fold tuning step above, run on the outer training data only. The analysis table had one row per patient, so no patient appeared in training and test data at the same time. Each patient’s out-of-fold predictions were averaged over the 10 repeats, and these pooled out-of-fold predictions were used for all summaries of discrimination, calibration and clinical utility. The primary metric was the area under the receiver operating characteristic curve (AUC), with a 95% confidence interval from a patient-level percentile bootstrap of the pooled out-of-fold predictions (2000 resamples). We also report the fold-to-fold standard deviation of the AUC, the area under the precision–recall curve, the Brier score and log-loss. Differences in AUC between the primary model and each comparator were computed fold by fold on identical folds and summarised with the Nadeau–Bengio corrected resampled t-interval [24], which accounts for the overlap of training sets in repeated cross-validation; p-values across comparators were adjusted with the Benjamini–Hochberg procedure [25].

Two further analyses checked the primary estimate. First, the optimism of the apparent AUC of the final model was estimated with Harrell’s bootstrap [26] (300 resamples), repeating the complete modelling strategy, including tuning, in every resample. Second, a label-permutation test [27] compared the cross-validated AUC (5-fold, regularisation fixed at the final value, mean over three fold assignments) with its distribution under 500 random permutations of the outcome, with p = (1 + b)/(1 + m), where b is the number of permutations reaching the observed AUC and m the number of permutations [28].

### Calibration and clinical utility

Calibration [29] was assessed with the calibration slope (ideal 1) and calibration-in-the-large (ideal 0), each with a 95% patient-level bootstrap interval (1000 resamples), and with a calibration plot showing a locally weighted (LOWESS) curve [30] and observed proportions in six quantile groups with Wilson intervals [31]. To express performance in terms of patients, we divided the pooled out-of-fold scores into tertiles (lower, middle and higher risk) and report the observed CKD proportion in each. In an illustrative testing-prioritisation scenario, we computed the share of all CKD cases found if only the highest-scoring 10% to 60% of patients were sent for laboratory testing, with bootstrap intervals, and the lift relative to testing the same number of patients at random. Decision curve analysis [32] compared the net benefit of testing according to the model with testing everyone or no one over threshold probabilities from 0.05 to 0.90. All probabilities refer to the study sample, in which 46% of patients had CKD.

### Sample size

The sample size was fixed by the available data. Its adequacy for developing a 20-parameter model was assessed retrospectively with the criteria of Riley and colleagues [14]: expected shrinkage of at least 0.90, an optimism of at most 0.05 in the Nagelkerke R², and estimation of the overall outcome proportion within ±0.05. The anticipated Cox–Snell R² was taken from the cross-validated predictions of the primary model, a deliberately conservative choice. The implementation was checked against the pmsampsize package. Events per variable are reported for orientation only.

### Predictor effects and plausibility audit

The final ridge model was fitted to all 400 patients with the regularisation strength selected by cross-validation, and its full equation is reported with the results. Its coefficients are reported as odds ratios per one standard deviation of each predictor, so that binary and continuous predictors share one scale, with 95% percentile intervals and the proportion of resamples with the same sign (sign agreement) from 2000 bootstrap refits. Permutation importance, the mean fall in AUC when one predictor is permuted, was computed on the held-out folds of the primary analysis (five permutations per fold). For a plausibility audit, the expected direction of association of each predictor with CKD was specified in the analysis code before the model was fitted, based on clinical knowledge (Table A in S1 Appendix). An effect was classified as supported when its interval excluded 1, and supported effects were compared with the expected direction. For readers who need inference on individual coefficients, an unpenalised multivariable logistic regression with Wald confidence intervals and Benjamini–Hochberg-adjusted p-values is also reported.

### Subgroups and sensitivity analyses

To check whether performance differed between groups of patients, discrimination (AUC with bootstrap interval) and calibration (ratio of observed to expected cases) were examined within subgroups of gender, age tertile, body-mass index category (below 18.5, 18.5–22.9, 23–24.9 and at least 25 kg/m², the cut-offs proposed for Asian populations [33]) and diabetes duration at the start of the record (1 year, 2–7 years, 8 years or more). Subgroups with fewer than 20 patients were not analysed.

Three sensitivity analyses repeated the nested cross-validation: after removing patients with an outlying value of age, diabetes duration, body-mass index or calorie intake (Tukey’s 1.5 × interquartile range rule [34]); with cubic spline terms (four knots) for the four continuous predictors, to check the linearity assumption; and with a different random assignment of folds. A feature-group ablation evaluated four clinical domains (demographics and body; diabetes history and treatment; comorbidity, urinary symptoms and pain-killer use; lifestyle and substance use) alone and by leaving each out of the full model, with the regularisation strength fixed at the final value and the same 50 folds.

### Secondary questions

#### Diabetes management pattern

Diet adherence, oral medicine and insulin together describe how a patient’s diabetes was managed, so they are not independent. We described the CKD proportion in each of the eight possible combinations (Wilson intervals; Monte Carlo permutation test of the chi-square statistic, 10,000 permutations) and tested whether seven pattern indicators, or three pairwise interaction terms, improved the cross-validated AUC beyond the three separate indicators (ridge model with fixed regularisation, same folds, Nadeau–Bengio intervals). We also compared how often the pattern changed during years 1 to 4 between patients later diagnosed with CKD and patients never diagnosed, restricted to patients still free of CKD at year 4 (Fisher’s exact test).

#### Value of the yearly history

For each time-varying feature we computed the share of year-to-year transitions in which its value changed. Landmark analyses [35] at record years 3, 5 and 7 included the patients still free of CKD at the landmark and classified whether CKD was diagnosed later in the record. Four models were compared on identical folds (stratified 5-fold cross-validation repeated 5 times): (A) ridge regression on the predictor values at the landmark; (B) ridge regression on the same values plus trajectory summaries (mean over years 1 to the landmark, change since year 1 and number of changes) for features that changed in at least 0.5% of transitions; (C) a neural network with one hidden layer of 16 units on the inputs of B; and (D) gradient boosting on the inputs of B. Differences from model A were summarised with Nadeau– Bengio intervals. Recurrent networks were not fitted, because their input would have been the same nearly constant series.

#### Sampling design

To describe when diagnoses fall within the record, we compared the share of first diagnoses in the final (collection) year with the 1 in 9 expected if diagnoses were spread evenly over years 2 to 10 (exact binomial test), and tested whether the remaining diagnoses were spread evenly over years 2 to 9 (chi-square test). Because a sample with 46% CKD is enriched for cases, we examined the consequences in a semi-synthetic simulation anchored to the cohort [36,37]. Predictor rows were resampled from the 400 patients, and outcomes were generated from the fitted ridge model with the intercept set to give a population prevalence of 10%, 20% or 30%. Models were trained either on a case-enriched sample of the cohort’s composition (185 cases and 215 controls) or on a random sample of 400, and the case-enriched models were also evaluated after prior correction, which adds log[π/(1−π)] − log[ȳ/(1−ȳ)] to the predicted log-odds, where π is the population prevalence and ȳ the sample proportion [37]. Each design was replicated 300 times per prevalence and evaluated on 20,000 independent simulated patients (AUC, mean predicted risk, calibration-in-the-large and slope).

#### Reliability of single train/test splits

Many published models are evaluated on one random split. We quantified the consequences in four ways [12]. (a) The ridge, random forest and gradient boosting models, with fixed hyper-parameters, were evaluated on 300 random stratified 80/20 splits of the cohort; the expected result of reporting the best of k random seeds was obtained by resampling the observed distribution (5000 draws). (b) In a simulation with 10 standard-normal predictors, an outcome prevalence of 46% and a known true AUC of 0.65, 0.75 or 0.85, 100 datasets were generated for each sample size from 100 to 1600. In each dataset we compared the estimate from a single 80/20 split, and the best of 3, 5 or 10 splits, with the estimate from 5-fold cross-validation repeated three times; errors were measured against the true AUC of the evaluated logistic regression model(s) on 8000 independent simulated patients. (c) A learning curve repeated 5-fold cross-validation of the ridge model on stratified subsamples of 100 to 400 patients. (d) The split experiment was repeated with logistic regression on seven public clinical datasets from OpenML [38].

### Statistical methods and software

Characteristics of patients with and without CKD were compared with the Mann–Whitney U test for continuous variables, Fisher’s exact test for binary variables and the chi-square test for job exertion level, and summarised with standardised mean differences [39]; p-values across the 20 comparisons were adjusted with the Benjamini–Hochberg procedure. Collinearity was checked with variance inflation factors. All tests were two-sided with α = 0.05; p-values of 0.001 or more are reported exactly and smaller values as p < 0.001.

Analyses were run in Python 3.13 with NumPy 2.1.3, pandas 2.2.3, scikit-learn 1.6.1 [40], SciPy 1.16.3, statsmodels 0.15.0 [41] and matplotlib 3.10.0. A master random seed (42) was used, from which each analysis derived its own independent random stream, so results do not depend on the order in which analyses are run. The code is available on GitHub (https://github.com/PanzerFaustAnan/diabetic-ckd-questionnaire-evaluation) and archived on Zenodo (version 1.0.0, https://doi.org/10.5281/zenodo.23758090) under the MIT licence [42].

### Ethics statement

Data collection was approved by the Ethical Review Committee of the Diabetic Association of Bangladesh (reference BADAS-ERC/EC/24/11), and the data-collection protocol was approved by BIRDEM General Hospital. All participants gave written informed consent before taking part [16]. The data were anonymised at the point of collection and no identifying information was recorded. The present study analyses only the published, de-identified dataset, and no additional ethical approval was sought.

## Results

### Participants and the design of the data

Of the 400 patients, 185 (46.3%) had a hospital diagnosis of CKD within their 10-year record. At the start of the record, patients who were later or already diagnosed differed from the others in 12 of the 20 characteristics after correction for multiple testing, but every difference was small to moderate (Table 1). The largest standardised differences were for urinary infection or urological issue (35.7% vs 12.6%), insulin use (54.6% vs 33.5%), diet adherence (74.6% vs 89.3%), regular pain-killer use (38.4% vs 21.9%), urinating properly, body-mass index and diabetes duration. Age did not differ (46.7 vs 46.1 years). Two differences ran against clinical expectation: hypertension was less common among patients with CKD (19.5% vs 31.6%) and regular walking more common (70.8% vs 58.1%). Collinearity was negligible (largest variance inflation factor 1.56; Table B in S1 Appendix).

**Table 1.** Characteristics at the start of the record by CKD status (n = 400).

| **Characteristic** | **No CKD (n = 215)** | **CKD (n = 185)** | **SMD** | **p** | **q** |
|---|---|---|---|---|---|
| Gender, male | 71 (33.0%) | 74 (40.0%) | +0.15 | 0.175 | 0.233 |
| Job exertion level, intermediate or heavy | 36 (16.7%) | 45 (24.3%) | +0.19 | 0.057 | 0.085 |
| Family history of diabetes | 86 (40.0%) | 83 (44.9%) | +0.10 | 0.361 | 0.380 |
| Diabetes duration (years), mean ± SD | 3.9 ± 5.4 | 5.8 ± 6.1 | +0.33 | <0.001 | <0.001 |
| Age (years), mean ± SD | 46.1 ± 10.5 | 46.7 ± 10.7 | +0.06 | 0.852 | 0.852 |
| Body-mass index (kg/m²), mean ± SD | 24.0 ± 3.9 | 25.2 ± 3.6 | +0.33 | <0.001 | 0.002 |
| Follows suggested diet | 192 (89.3%) | 138 (74.6%) | −0.39 | <0.001 | <0.001 |
| Takes oral diabetes medicine | 168 (78.1%) | 134 (72.4%) | −0.13 | 0.201 | 0.251 |
| Takes insulin | 72 (33.5%) | 101 (54.6%) | +0.44 | <0.001 | <0.001 |
| Hypertension | 68 (31.6%) | 36 (19.5%) | −0.28 | 0.006 | 0.012 |
| Heart disease | 35 (16.3%) | 53 (28.6%) | +0.30 | 0.004 | 0.009 |
| Sufficient sleep (7–9 h) | 139 (64.7%) | 96 (51.9%) | −0.26 | 0.011 | 0.018 |
| Sufficient water intake (>2 L/day) | 199 (92.6%) | 165 (89.2%) | −0.12 | 0.294 | 0.326 |
| Smoking | 23 (10.7%) | 32 (17.3%) | +0.19 | 0.060 | 0.085 |
| Zarda / betel leaf use | 47 (21.9%) | 50 (27.0%) | +0.12 | 0.244 | 0.287 |
| Walks regularly (30+ min/day) | 125 (58.1%) | 131 (70.8%) | +0.27 | 0.009 | 0.017 |
| Urinates properly | 196 (91.2%) | 147 (79.5%) | −0.34 | <0.001 | 0.003 |
| Urinary infection / urological issue | 27 (12.6%) | 66 (35.7%) | +0.56 | <0.001 | <0.001 |
| Regular pain-killer use | 47 (21.9%) | 71 (38.4%) | +0.37 | <0.001 | 0.002 |
| Calorie intake (kcal/day), mean ± SD | 1437.1 ± 238.4 | 1473.0 ± 174.5 | +0.17 | 0.004 | 0.009 |

Values are n (%) unless stated otherwise. SMD, standardised mean difference (CKD minus no CKD). p, Mann–Whitney U test (continuous), Fisher's exact test (binary) or chi-square test (job exertion, three levels); q, Benjamini–Hochberg-adjusted p-value across the 20 comparisons.

The first CKD diagnosis fell in the final (collection) year for 127 of the 185 patients with CKD (68.6%, 95% CI 61.6–74.9%), against 11.1% expected if diagnoses were spread evenly over the record (p < 0.001; Fig 1A). The remaining 58 diagnoses were spread over years 2 to 9, though not evenly (p = 0.001), with few in the earliest years. For 217 patients (54.3%) the record began in the first year after the diabetes diagnosis (Fig 1B). This pattern indicates that patients with CKD were mostly recruited around the time of their diagnosis, which makes the sample case-enriched and means that the timing of diagnosis within the record partly reflects recruitment.

**Fig 1.**
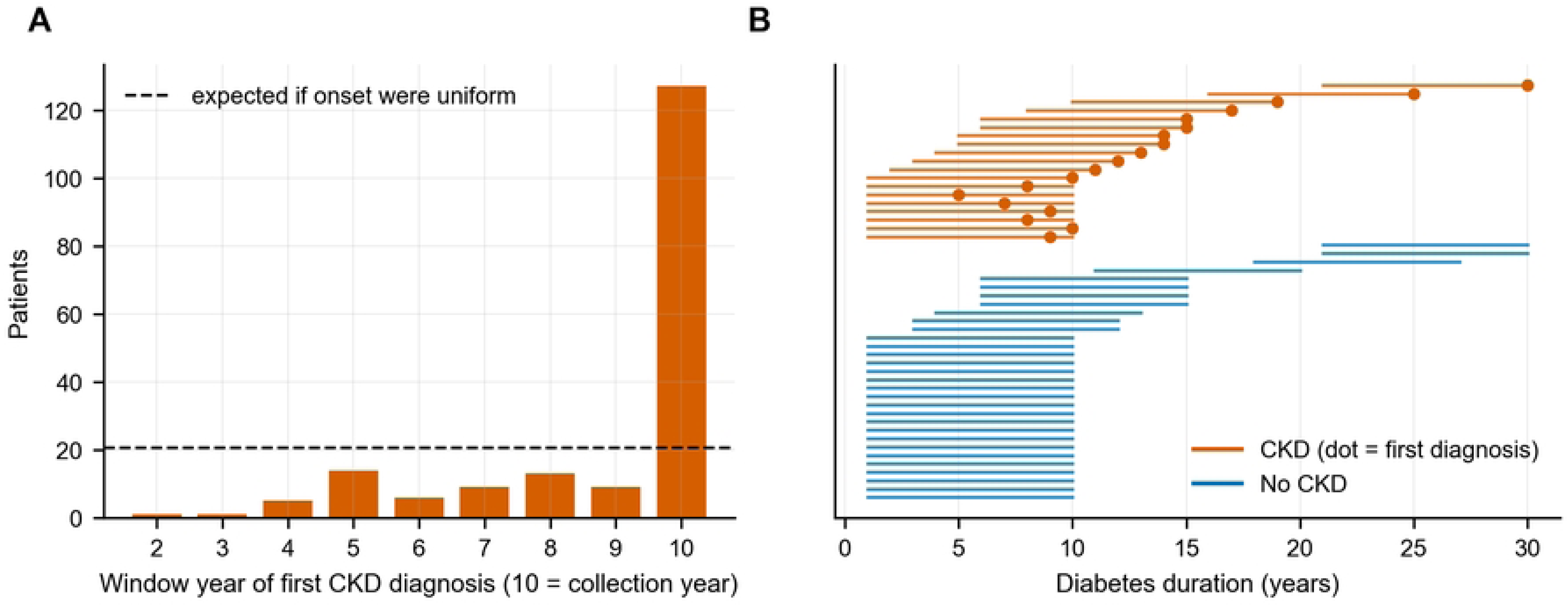
Timing of CKD diagnoses within the 10-year records. (A) Record year of the first CKD diagnosis among the 185 patients with CKD (year 10 is the collection year); the dashed line is the count expected per year if diagnoses were spread evenly over years 2–10. (B) Records of 50 randomly selected patients on the diabetes-duration scale; dots mark the first CKD diagnosis.

## Discrimination

The pre-specified ridge model had an AUC of 0.751 (95% CI 0.701–0.795) under nested cross-validation, with a fold-to-fold standard deviation of 0.047 (Table 2, Fig 2). The lasso, random forest and gradient boosting models reached AUCs of 0.748 to 0.764, and none differed detectably from the ridge model, with paired differences of −0.005 to +0.006 and intervals of up to ±0.045 (Table 3). The ridge model clearly outperformed a model using age and diabetes duration only (AUC 0.562; difference 0.159, 95% CI 0.082–0.236) and the prevalence-only model. The area under the precision–recall curve was 0.724 (0.658–0.788), against a chance level of 0.463.

**Fig 2.**
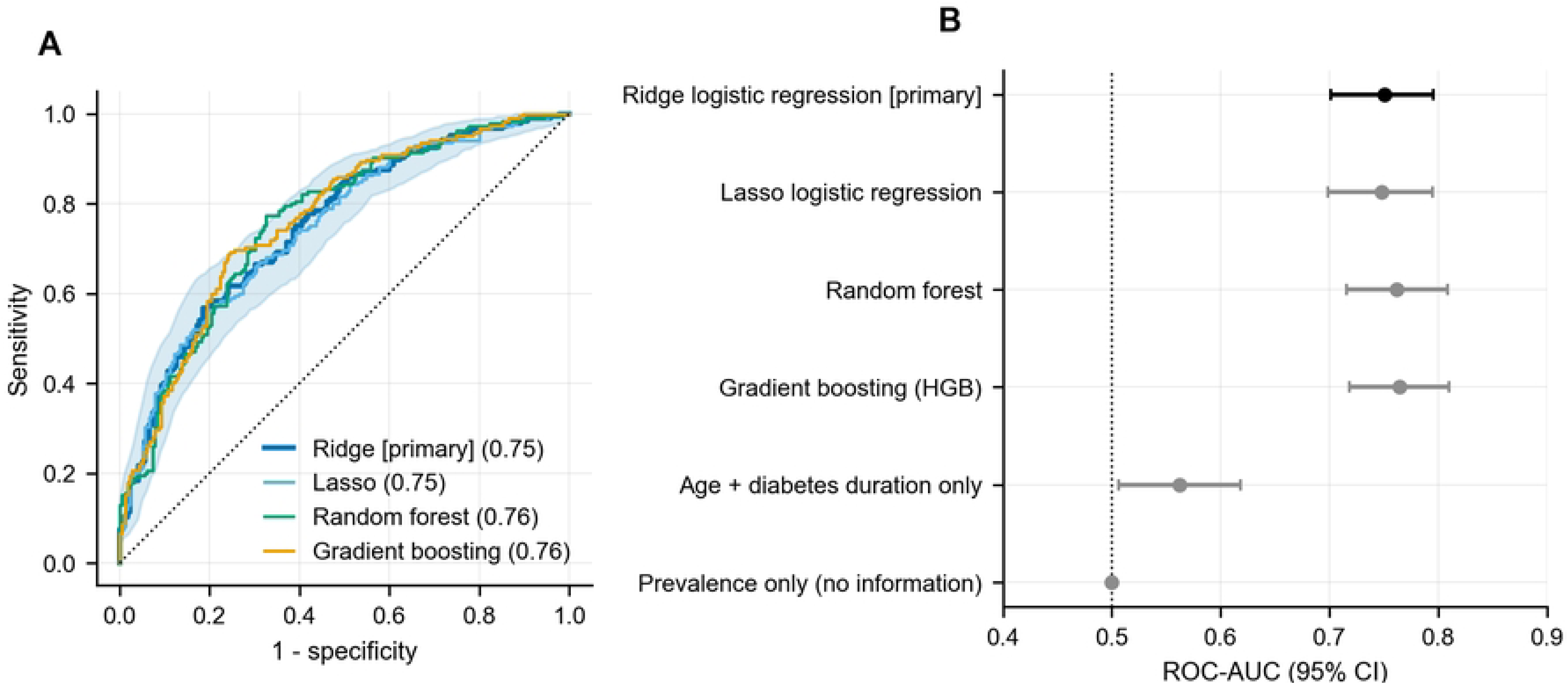
Discrimination under nested repeated cross-validation. (A) Receiver operating characteristic curves from pooled out-of-fold predictions; the shaded band is the 95% bootstrap band for the primary ridge model, and the AUC is given in brackets. (B) AUC with 95% patient-level bootstrap confidence intervals for the primary model, comparators and baselines (n = 400).

**Table 2.** Discrimination and accuracy under nested repeated cross-validation.

| **Model** | **AUC (95% CI)** | **AUC, fold mean ± SD** | **PR-AUC (95% CI)** | **Brier score** | **Log-loss** |
|---|---|---|---|---|---|
| Ridge logistic regression (primary) | 0.751 (0.701 to 0.795) | 0.748 ± 0.047 | 0.724 (0.658 to 0.788) | 0.204 | 0.595 |
| Lasso logistic regression | 0.748 (0.699 to 0.795) | 0.743 ± 0.046 | 0.723 (0.658 to 0.787) | 0.205 | 0.597 |
| Random forest | 0.762 (0.716 to 0.808) | 0.752 ± 0.045 | 0.732 (0.667 to 0.793) | 0.203 | 0.593 |
| Gradient boosting | 0.764 (0.718 to 0.810) | 0.753 ± 0.047 | 0.731 (0.668 to 0.791) | 0.202 | 0.590 |
| Age + diabetes duration only | 0.562 (0.506 to 0.619) | 0.589 ± 0.057 | 0.521 (0.457 to 0.602) | 0.245 | 0.684 |
| Prevalence only (no information) | 0.500 (0.500 to 0.500) | 0.500 ± 0.000 | 0.463 (0.412 to 0.510) | 0.249 | 0.690 |

400 patients; outer stratified 5-fold cross-validation repeated 10 times with 3-fold inner tuning. AUC and PR-AUC are computed from pooled out-of-fold predictions with 95% CIs from a patient-level bootstrap (2000 resamples); Brier score and log-loss are means over the 50 test folds. AUC, area under the receiver operating characteristic curve; PR-AUC, area under the precision–recall curve (chance level 0.463).

**Table 3.**
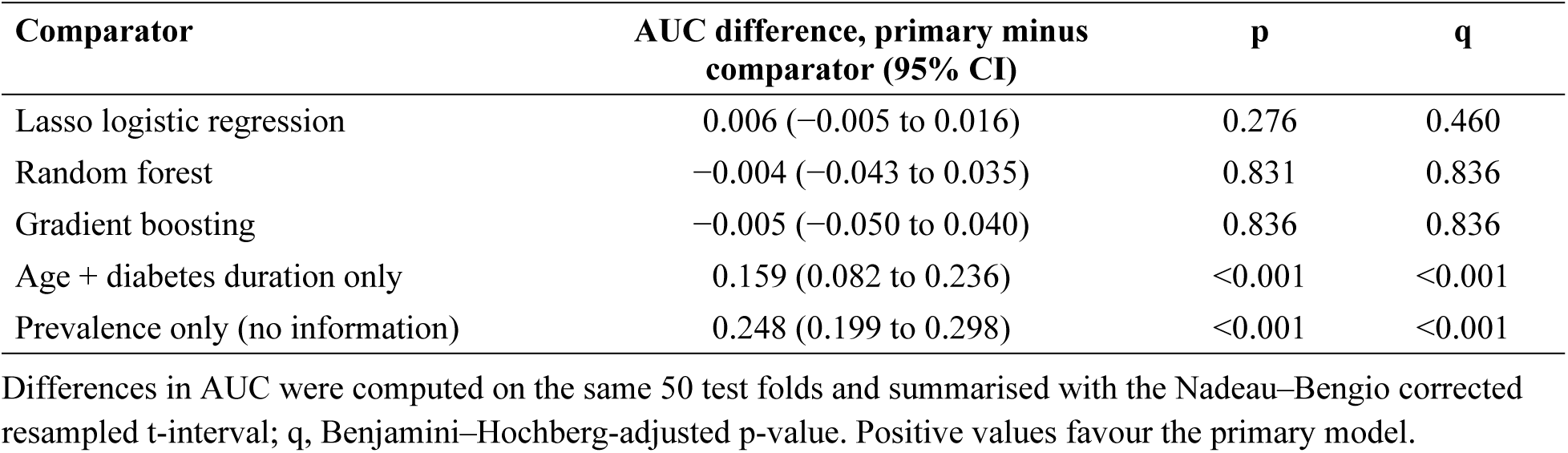
Paired comparison of the primary ridge model with each comparator.

| **Comparator** | **AUC difference, primary minus comparator (95% CI)** | **p** | **q** |
|---|---|---|---|
| Lasso logistic regression | 0.006 (−0.005 to 0.016) | 0.276 | 0.460 |
| Random forest | −0.004 (−0.043 to 0.035) | 0.831 | 0.836 |
| Gradient boosting | −0.005 (−0.050 to 0.040) | 0.836 | 0.836 |
| Age + diabetes duration only | 0.159 (0.082 to 0.236) | <0.001 | <0.001 |
| Prevalence only (no information) | 0.248 (0.199 to 0.298) | <0.001 | <0.001 |

Differences in AUC were computed on the same 50 test folds and summarised with the Nadeau–Bengio corrected resampled t-interval; q, Benjamini–Hochberg-adjusted p-value. Positive values favour the primary model.

Two further checks agreed with the cross-validated estimate. The apparent AUC of the final model (selected penalty C = 0.1) was 0.800, the bootstrap estimate of optimism was 0.043, and the optimism-corrected AUC was 0.757. In the permutation test, the cross-validated AUC of 0.747 exceeded every one of 500 AUCs obtained with shuffled outcomes (null mean 0.502, largest 0.627; p = 0.002), so the features carry real information about CKD status.

The sample was smaller than recommended for developing a 20-parameter model (Table 4). With the cross-validated Cox–Snell R² of 0.182, the criterion of small overfitting required 883 patients, more than twice the 400 available; the precision criterion for the overall proportion (382 patients) was met. There were 9.25 events per candidate parameter.

**Table 4.** Retrospective assessment of sample size against the criteria of Riley and colleagues.

| **Criterion** | **Minimum number of patients** | **Available / required** |
|---|---|---|
| 1. Expected shrinkage ≥ 0.90 | 883 | 0.45 |
| 2. Optimism in Nagelkerke R² ≤ 0.05 | 473 | 0.85 |
| 3. Overall proportion estimated within ±0.05 | 382 | 1.05 |
| Required (largest of 1–3) | 883 | 0.45 |
| Available | 400 | 1.00 |

20 candidate parameters; outcome proportion 0.463; anticipated Cox–Snell R² 0.182 (from cross-validated predictions); 185 events, 9.25 events per candidate parameter.

### Calibration and clinical utility

Within the sample, predicted probabilities agreed with observed proportions: the calibration slope was 1.06 (95% CI 0.79–1.33) and calibration-in-the-large −0.003 (−0.227 to 0.209) (Fig 3A). The Brier score was 0.202, against 0.249 for the prevalence-only model. In the lower, middle and higher thirds of predicted probability, 20.9% (14.9–28.5%), 45.9% (37.6–54.3%) and 72.2% (64.0–79.1%) of patients had CKD (Fig 3B; Table C in S1 Appendix). If laboratory testing were limited to the highest-scoring 30% of patients, 48% (43–53%) of all CKD cases would be found, 1.6 times as many as by testing the same number of patients at random (Table 5, Fig 3C). In decision curve analysis, testing by the model gave a higher net benefit than testing everyone at threshold probabilities above about 0.2, for example 0.206 against 0.104 at a threshold of 0.4 (Fig 3D; Table D in S1 Appendix). These probabilities and net benefits apply to a population with 46% CKD.

**Fig 3.**
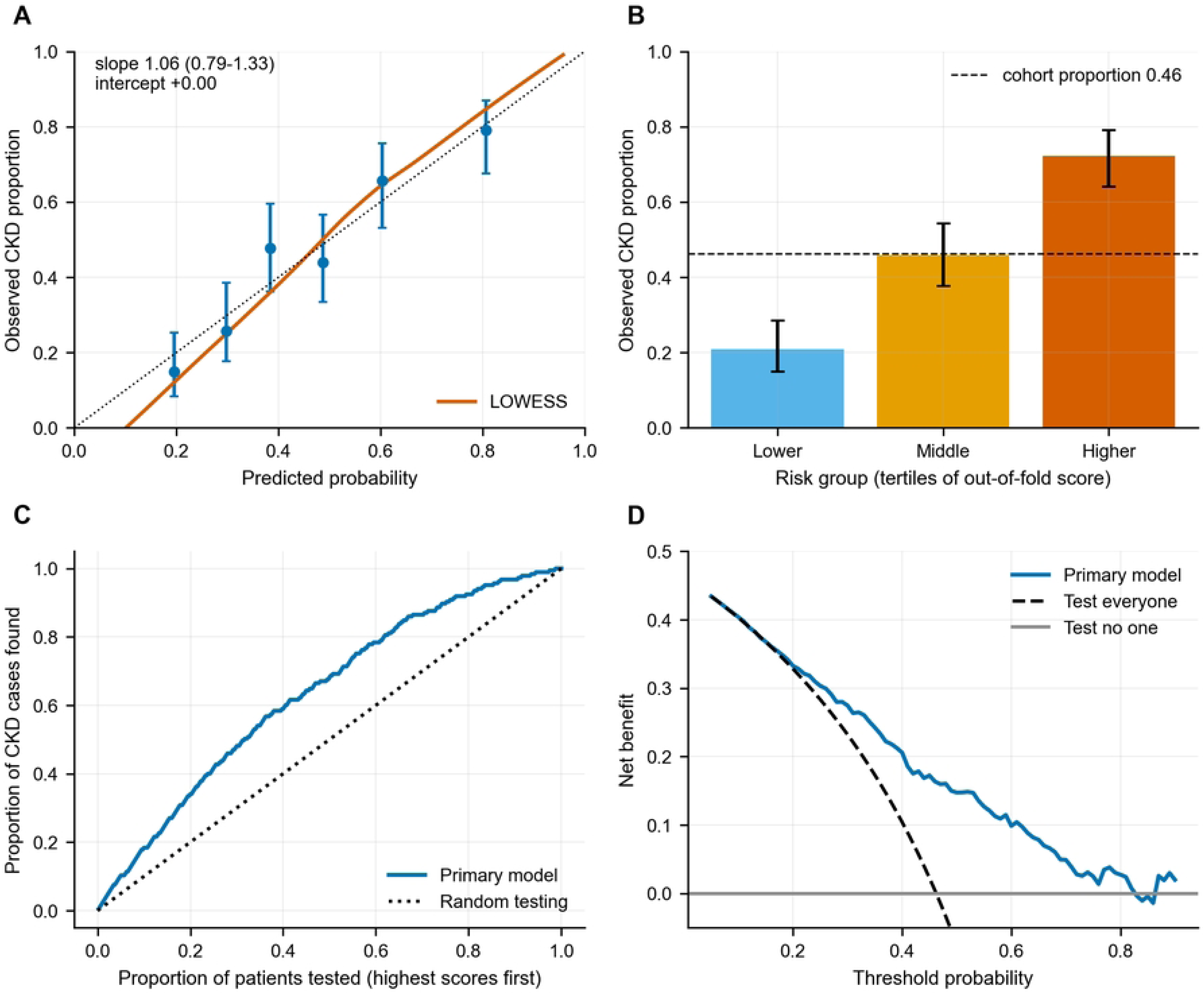
Calibration and clinical utility of the primary model. (A) Calibration plot with a LOWESS curve and observed proportions in six quantile groups with 95% Wilson intervals; the dotted line is perfect calibration. (B) Observed CKD proportion (95% Wilson interval) in tertiles of predicted probability; the dashed line is the overall proportion. (C) Cumulative share of CKD cases found when patients are tested in order of decreasing score; the dotted line is random testing. (D) Decision curve: net benefit of testing by the model, testing everyone and testing no one. All probabilities refer to a sample with 46% CKD.

**Table 5.** Testing-prioritisation scenario: CKD cases found when only the highest-scoring patients are tested.

| **Patients tested (highest scores first)** | **Share of CKD cases found (95% CI)** | **CKD proportion among those tested** | **Lift over random testing** |
|---|---|---|---|
| 10% | 0.18 (0.15 to 0.21) | 0.85 | 1.84 |
| 20% | 0.34 (0.30 to 0.38) | 0.79 | 1.70 |
| 30% | 0.48 (0.43 to 0.53) | 0.74 | 1.60 |
| 40% | 0.59 (0.55 to 0.65) | 0.68 | 1.47 |
| 50% | 0.68 (0.63 to 0.74) | 0.63 | 1.36 |
| 60% | 0.78 (0.73 to 0.84) | 0.60 | 1.31 |

Based on pooled out-of-fold predictions of the primary model in a sample with 46% CKD; 95% CIs from a patient-level bootstrap (1000 resamples). Lift is the share of cases found divided by the share of patients tested.

Discrimination varied across subgroups, from 0.64 (95% CI 0.52–0.75) in patients with a body-mass index of 23–24.9 kg/m² to 0.82 (0.75–0.88) in the youngest age tertile, and was 0.77 (0.72– 0.83) in women and 0.70 (0.62–0.78) in men (Fig A and Table E in S1 Appendix). The intervals overlapped widely, and the ratio of observed to expected cases ranged from 0.86 to 1.18, so these differences should be treated as hypotheses.

### Predictor effects and plausibility audit

Eight predictors had bootstrap intervals that excluded an odds ratio of 1, and their signs were stable in at least 99.75% of resamples (Table 6, Fig 4). Six agreed with clinical expectation: body-mass index (odds ratio per standard deviation 1.49, 95% CI 1.23–1.87), urinary infection or urological issue (1.45, 1.18–1.83), insulin use (1.38, 1.14–1.71), diabetes duration (1.36, 1.10– 1.68) and regular pain-killer use (1.33, 1.09–1.66) were associated with higher odds of CKD, and diet adherence (0.71, 0.56–0.87) with lower odds. Two contradicted it: regular walking was associated with higher odds (1.44, 1.18–1.78) and hypertension with lower odds (0.73, 0.59– 0.88). Permuting urinary infection, body-mass index, insulin use, diet adherence or regular walking reduced the AUC most (by 0.025 to 0.040). The full model equation is given in Table F in S1 Appendix. An unpenalised multivariable logistic regression identified the same eight predictors (all with Benjamini–Hochberg-adjusted p < 0.05; Table G in S1 Appendix).

**Fig 4.**
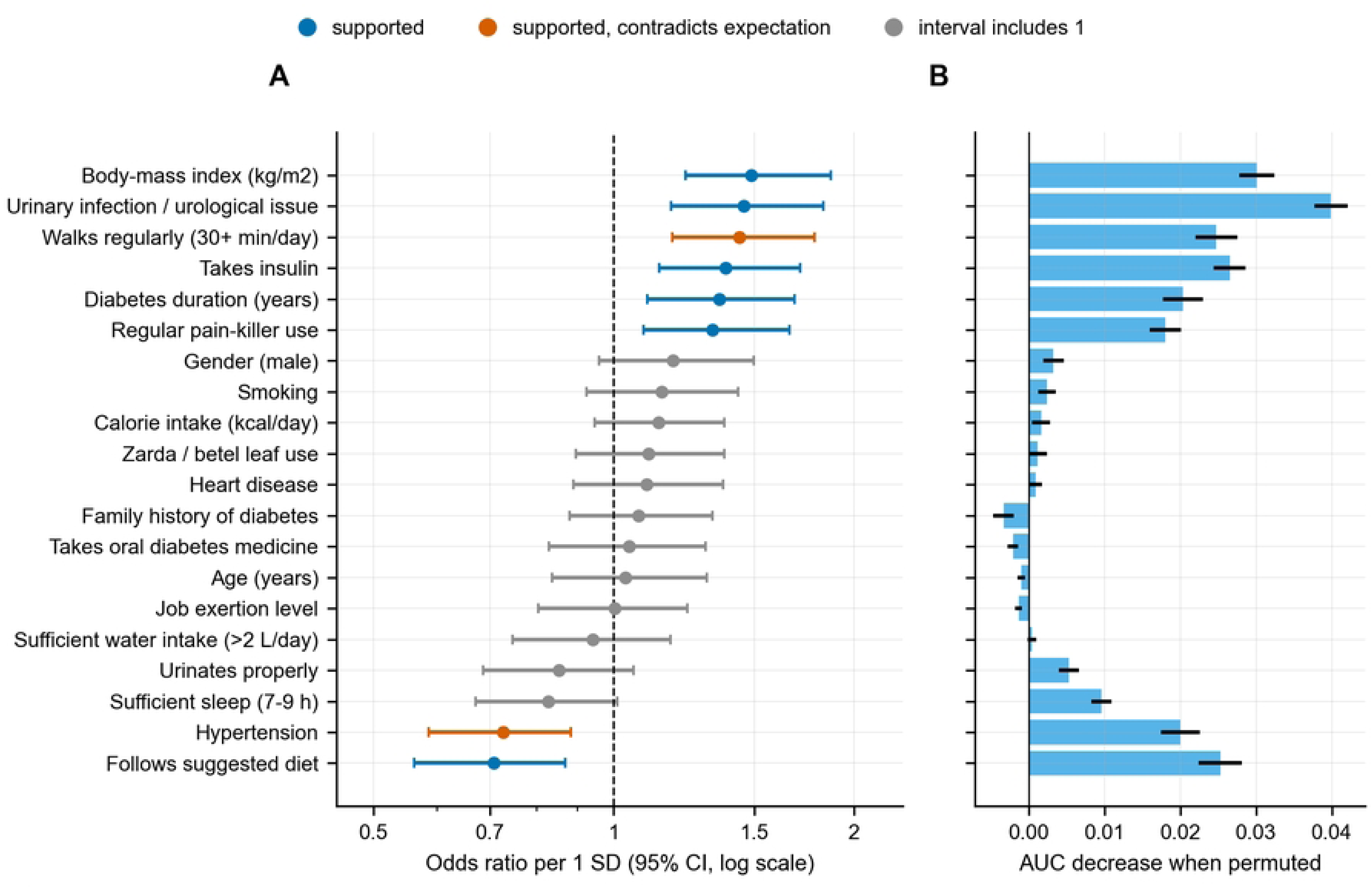
Predictor effects in the final ridge model. (A) Odds ratio per one standard deviation with 95% bootstrap intervals, coloured by the plausibility audit. (B) Mean fall in AUC when the predictor is permuted in held-out folds (error bars, standard error over 50 folds), in the same order as panel A. Associations are cross-sectional and not causal.

**Table 6.** Predictor effects in the final ridge model.

| **Predictor** | **Odds ratio per SD (95% CI)** | **Sign agreement** | **Permutation importance** | **Plausibility audit** |
|---|---|---|---|---|
| Body-mass index (kg/m²) | 1.49 (1.23 to 1.87) | 100% | 0.030 | Supported, as expected |
| Urinary infection / urological issue | 1.45 (1.18 to 1.83) | 100% | 0.040 | Supported, as expected |
| Walks regularly (30+ min/day) | 1.44 (1.18 to 1.78) | 100% | 0.025 | Supported, contradicts expectation |
| Takes insulin | 1.38 (1.14 to 1.71) | 100% | 0.027 | Supported, as expected |
| Diabetes duration (years) | 1.36 (1.10 to 1.68) | 100% | 0.020 | Supported, as expected |
| Regular pain-killer use | 1.33 (1.09 to 1.66) | 100% | 0.018 | Supported, as expected |
| Gender (male) | 1.19 (0.96 to 1.50) | 94% | 0.003 | Interval includes 1 |
| Smoking | 1.15 (0.92 to 1.43) | 89% | 0.002 | Interval includes 1 |
| Calorie intake (kcal/day) | 1.14 (0.95 to 1.38) | 91% | 0.002 | Interval includes 1 |
| Zarda / betel leaf use | 1.11 (0.90 to 1.38) | 85% | 0.001 | Interval includes 1 |
| Heart disease | 1.10 (0.89 to 1.37) | 82% | 0.001 | Interval includes 1 |
| Family history of diabetes | 1.07 (0.88 to 1.33) | 78% | -0.003 | Interval includes 1 |
| Takes oral diabetes medicine | 1.05 (0.83 to 1.30) | 65% | -0.002 | Interval includes 1 |
| Age (years) | 1.03 (0.84 to 1.31) | 62% | -0.001 | Interval includes 1 |
| Job exertion level | 1.00 (0.80 to 1.24) | 50% | -0.001 | Interval includes 1 |
| Sufficient water intake (>2 L/day) | 0.94 (0.75 to 1.18) | 71% | 0.000 | Interval includes 1 |
| Urinates properly | 0.85 (0.69 to 1.06) | 92% | 0.005 | Interval includes 1 |
| Sufficient sleep (7–9 h) | 0.83 (0.67 to 1.01) | 97% | 0.010 | Interval includes 1 |
| Hypertension | 0.73 (0.59 to 0.88) | 100% | 0.020 | Supported, contradicts expectation |
| Follows suggested diet | 0.71 (0.56 to 0.87) | 100% | 0.025 | Supported, as expected |

Ridge logistic regression fitted to all 400 patients (C = 0.1). Odds ratios per one standard deviation of each predictor with 95% percentile intervals from 2000 bootstrap refits; sign agreement is the share of refits with the same sign. Permutation importance is the mean fall in AUC when the predictor is permuted in held-out folds. Associations are cross-sectional and not causal.

The results were robust to analytic choices (Table H in S1 Appendix). The AUC was 0.769 after removing 55 patients with an outlying continuous value, 0.768 with spline terms (difference from the primary model +0.017, 95% CI −0.019 to +0.054) and 0.754 with a different assignment of folds. In the feature-group ablation, removing the comorbidity, urinary and pain-killer group reduced the AUC by 0.051 (0.021–0.081) and removing the diabetes history and treatment group by 0.034 (0.006–0.063), whereas removing the demographic or lifestyle groups changed it by less than 0.015 (Table I in S1 Appendix).

### Secondary questions

The proportion with CKD differed across the eight management patterns (p < 0.001), from 32% among patients following diet and oral medicine (n = 173) to 80% among those using insulin only (n = 15) (Fig B and Table J in S1 Appendix). Adding pattern indicators (difference −0.003, 95% CI −0.019 to +0.014) or pairwise interactions (+0.000, −0.006 to +0.007) did not change the AUC (Table K in S1 Appendix), so the pattern carries no information beyond the three separate indicators. During years 1 to 4, the management pattern changed less often in patients later diagnosed with CKD than in those never diagnosed (21.3% vs 35.3%, p = 0.003).

Within patients, 9 of the 14 time-varying features changed in fewer than 1% of year-to-year transitions; diet adherence changed most often (17.8% of transitions) (Fig C and Table L in S1 Appendix). Adding trajectory summaries to the landmark snapshot changed the AUC by +0.006 (95% CI −0.018 to +0.031) at year 3, +0.018 (−0.018 to +0.054) at year 5 and +0.033 (0.000 to +0.067) at year 7; neither a neural network nor gradient boosting on the same inputs differed detectably from the snapshot model at any landmark (Table M in S1 Appendix). The yearly history therefore added no reliable information beyond the current profile.

### Effect of case-enriched sampling

In the simulation anchored to the cohort, models trained on a case-enriched sample had the same AUC whether or not their probabilities were corrected, but their predicted risks were far too high when applied to a population with lower prevalence (Fig 5; Table N in S1 Appendix). At a true prevalence of 10%, the mean predicted risk was 0.366 and calibration-in-the-large −2.05; at 30%, they were 0.426 and −0.71. Prior correction with the known prevalence removed this bias (mean predicted risks 0.101, 0.203 and 0.301 at prevalences of 10%, 20% and 30%), and models trained on a random sample of 400 were also unbiased on average but varied more between replicates.

**Fig 5.**
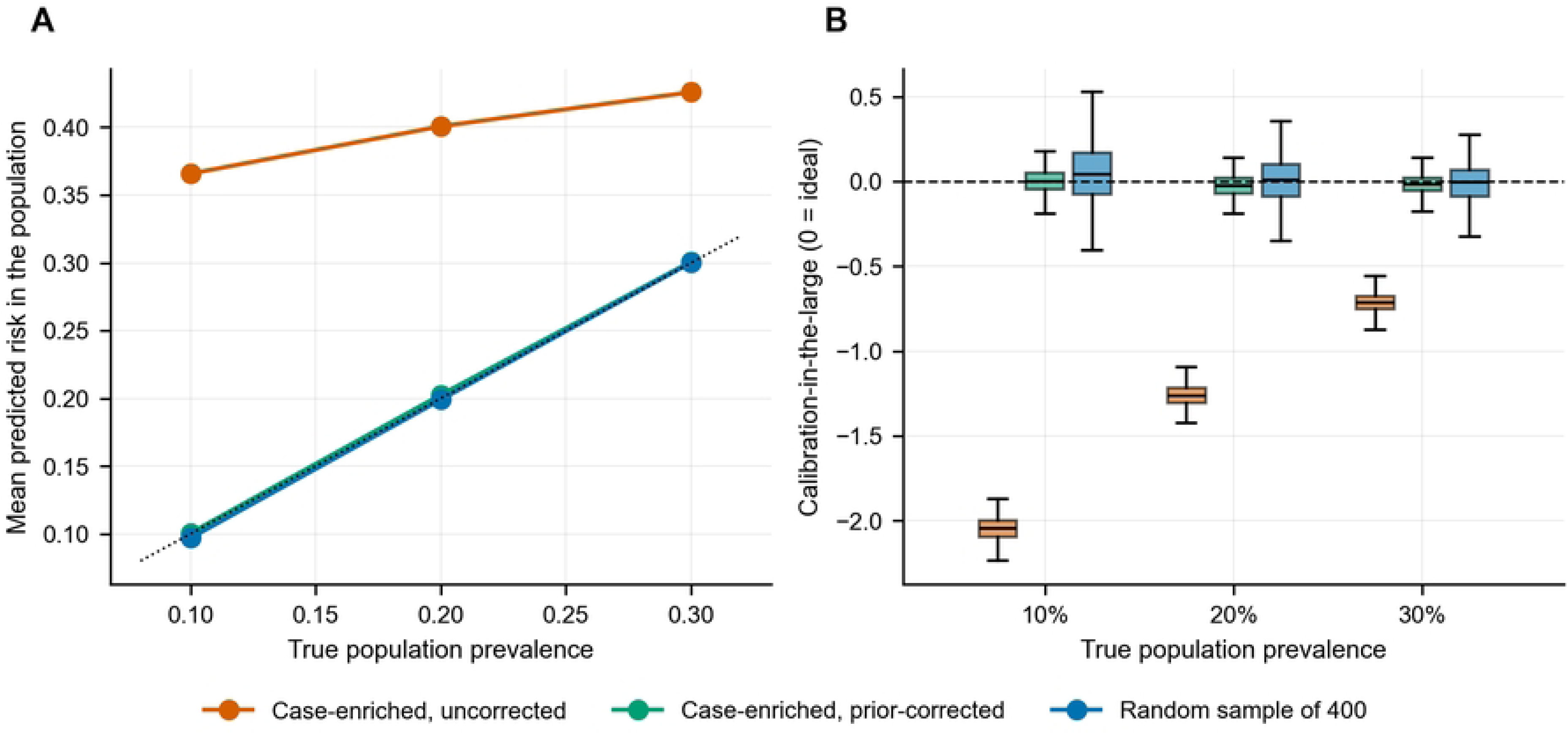
Effect of case-enriched sampling on predicted probabilities in simulation. (A) Mean predicted risk in the population against the true population prevalence for models trained on a case-enriched sample (185 cases and 215 controls) without and with prior correction, and on a random sample of 400; the dotted line is perfect agreement. (B) Calibration-in-the-large across 300 replicates per prevalence (0 is ideal).

### Reliability of single train/test splits

Over 300 random 80/20 splits of the cohort, the test AUC of the ridge model averaged 0.750 but ranged from 0.607 to 0.872, with 95% of splits between 0.657 and 0.836; the random forest and gradient boosting models varied in the same way (Table 7, Fig 6A). A researcher who tried ten random splits and reported the best would, on average, report an AUC of 0.820, 0.07 above the mean; with three or five splits the expected report was 0.789 and 0.802.

**Fig 6.**
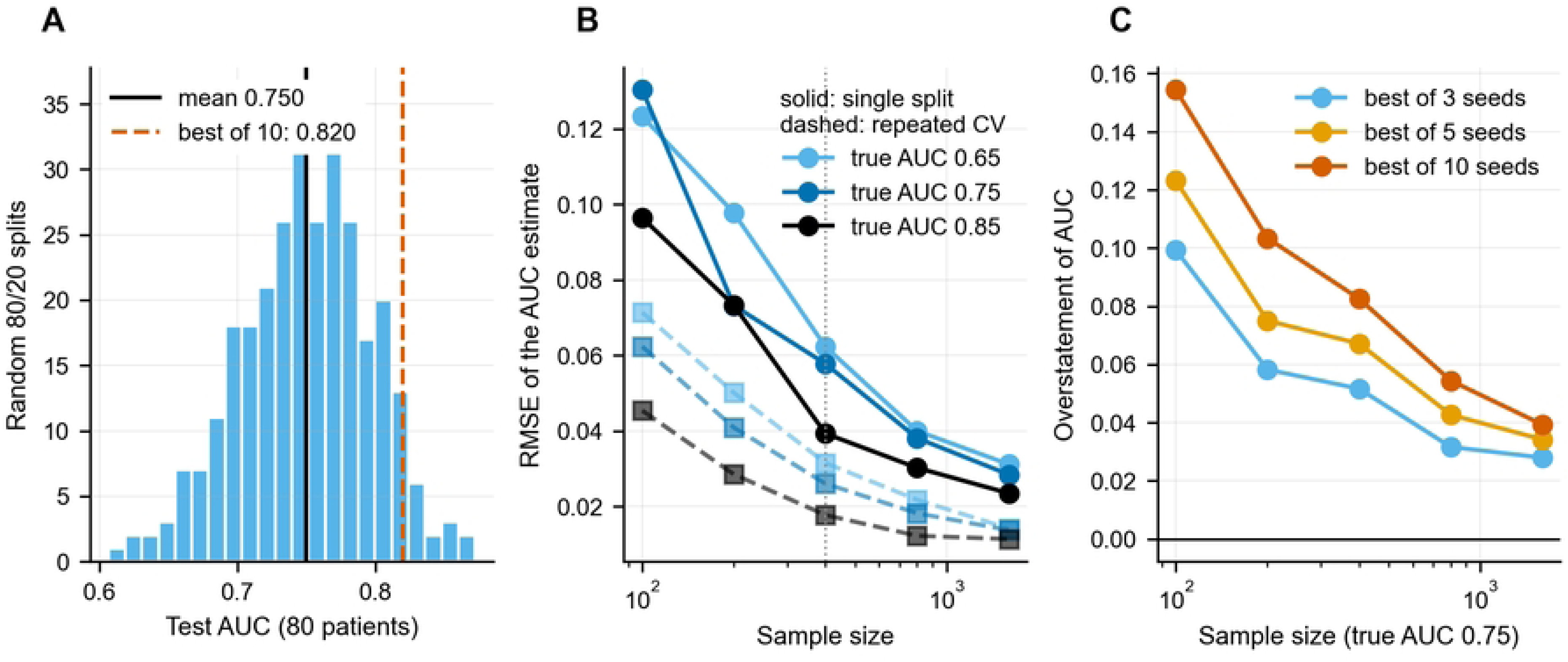
Reliability of single train/test splits. (A) Test AUC of the ridge model over 300 random 80/20 splits of the cohort; the solid line is the mean and the dashed line the expected value when the best of 10 splits is reported. (B) Root-mean-square error of the AUC estimate from a single 80/20 split (solid) and from repeated cross-validation (dashed) against sample size, in simulations with a known true AUC; the dotted line marks this cohort’s size. (C) Mean overstatement of the AUC when the best of k random splits is reported (true AUC 0.75).

**Table 7.** Test AUC over 300 random 80/20 splits of the cohort.

| Model | Mean test AUC | SD | Central 95% of splits | Range | Splits ≥ 0.05 above mean |
|---|---|---|---|---|---|
| Ridge logistic regression | 0.750 | 0.047 | 0.657–0.836 | 0.607–0.872 | 15.3% |
| Random forest | 0.759 | 0.048 | 0.654–0.847 | 0.607–0.872 | 13.3% |
| Gradient boosting | 0.750 | 0.050 | 0.646–0.835 | 0.588–0.865 | 16.0% |

Fixed hyper-parameters (no tuning), so only the split varies; each test set contains 80 patients.

The simulation with a known true AUC of 0.75 showed that this is a property of the sample size rather than of these data (Fig 6B and 6C; Table O in S1 Appendix). At 400 patients, the root-mean-square error of a single-split estimate was 0.058, against 0.026 for repeated cross-validation, and a single split overstated the true AUC by at least 0.05 in 27% of datasets. Reporting the best of ten splits overstated it by 0.083 on average at 400 patients and by 0.154 at 100 patients, and the overstatement fell only to 0.039 at 1600 patients. The learning curve of the ridge model was nearly flat, from 0.729 with 100 patients to 0.744 with 400 (Fig D in S1 Appendix). On seven public clinical datasets of 155 to 768 patients, the standard deviation of the single-split AUC ranged from 0.003 to 0.087 and followed the simulated relation with sample size (Fig E and Table P in S1 Appendix).

## Discussion

In 400 adults with long-standing diabetes attending a tertiary hospital in Dhaka, 20 non-invasive questionnaire features classified CKD status with an AUC of 0.75 (95% CI 0.70–0.80). The signal was real, robust to the choice of algorithm and to sensitivity analyses, and well calibrated within the sample, and ranking patients by the model would have found about half of the CKD cases among the highest-scoring 30%. The evidence is limited in three ways that the analysis makes explicit: the sample was less than half the size recommended for a model of this complexity, the features largely describe patients after diagnosis, and the case-enriched design means that the predicted probabilities do not apply to an ordinary clinic population without recalibration.

An AUC of 0.75 means that a randomly chosen patient with CKD received a higher score than a randomly chosen patient without CKD three times out of four. That is too weak for diagnosis but could be useful for deciding whom to test first where laboratory capacity is limited, which is the intended use of such a tool. Two conditions apply. First, the probabilities refer to a sample in which 46% of patients had CKD; our simulation shows that a model trained on such a sample would overstate risk in a population with 10% to 30% prevalence, and that the bias can be removed only if the target prevalence is known or the model is recalibrated locally [29,37]. Second, studies that compare cases identified at diagnosis with clearly unaffected controls tend to overestimate diagnostic accuracy [43]. Our simulation cannot capture this, because it assumes that the features relate to CKD in the same way in the sample and the population, so the AUC itself may be lower in consecutive patients.

The predictor associations should be read as cross-sectional. Most features were recorded as one value for the whole record or rarely changed, and for two thirds of the patients with CKD the diagnosis fell in the year of the interview. Several associations are therefore plausibly consequences of CKD or of its management rather than risk factors. Insulin use is a clear example: oral agents such as metformin are restricted at low kidney function, so patients are often moved to insulin as CKD develops [5]. Regular walking, which was associated with higher odds of CKD, is recommended to patients with diabetes and CKD [5], and patients may have reported the advice they had received after diagnosis. The positive association with regular pain-killer use agrees with the known nephrotoxicity of non-steroidal anti-inflammatory drugs [44], but the direction of the relation cannot be established here.

Hypertension, a recognised cause of CKD progression, was associated with lower odds of CKD. Two explanations are plausible and cannot be distinguished with these data. In the dataset, hypertension means sustained high blood pressure treated with medication, and angiotensin II receptor blockers and angiotensin-converting enzyme inhibitors, which slow the decline of kidney function in diabetes [45,46], are widely prescribed in Dhaka [47,48]. Treated hypertensive patients may therefore have been partly protected, although treatment more readily explains a weaker association than a reversed one. Second, eligibility required at least 10 years of diabetes and current attendance at the hospital; hypertensive patients whose kidney disease progressed fastest may have left this population through dialysis, transfer or death before they could be recruited, a form of selection bias that can reverse associations [49]. Because the drug class was not recorded, neither explanation can be tested. For these reasons the model should not be used to give advice about behaviour or treatment.

The ten-year yearly records added little. Most features hardly changed within patients, the gain from trajectory summaries was small and inconsistent across landmarks, and flexible models did not extract more from the same inputs. Studies that attribute added value to longitudinal modelling of such data should show it against a snapshot baseline evaluated on the same folds. Similarly, the diabetes management pattern was strongly associated with CKD, but the three separate indicators already carried this information.

The single-split experiments have implications beyond this dataset. Splitting the same 400 patients at random gave test AUCs from 0.61 to 0.87, and selecting the best of ten splits would have inflated the result by about 0.07. The simulation shows that error bars of this size are expected at a few hundred patients whatever the true accuracy, consistent with earlier work [12,13], and the public datasets show the same pattern. Published CKD models trained on small datasets and evaluated on a single split [8] should therefore be interpreted with caution. This dataset is no exception: one analysis treated its 4,000 patient-years as independent observations, split them at random and reported an accuracy of 0.94 [50]. Because about 90% of patient-years carry no CKD label, always predicting no CKD would score 0.90, and records of the same patient fall on both sides of such a split. Repeated, nested cross-validation with confidence intervals, a pre-specified primary model, and reporting of all analyses, as recommended by TRIPOD+AI [18], are inexpensive safeguards.

The strengths of this study are a pre-specified primary model and analysis plan, evaluation by nested repeated cross-validation at the patient level, confidence intervals for every performance measure, assessment of calibration and clinical utility, an explicit audit of the plausibility of the associations, simulations that separate design effects from features of the data, and public data and code that reproduce every number. The limitations are substantial. The data come from one tertiary centre and include only patients with at least 10 years of diabetes and a complete record book, who are likely to differ from the wider diabetic population. The sample was smaller than recommended, so the coefficients may be unstable even with penalisation. There was no external validation. Predictors were collected at a single structured interview and cross-checked against the record books, but lifestyle items relied on patient report, CKD status was visible to interviewers in the record books, several features were recorded as values for the whole record, and antihypertensive drug classes were not recorded. The model needs all 20 inputs and has no procedure for missing answers. Finally, the findings on subgroups were imprecise.

Before any practical use, the model would need external validation in consecutive patients with diabetes who complete the questionnaire before their CKD status is known, recalibration to the prevalence of the setting, and evaluation of its effect on testing decisions.

## Conclusions

Twenty non-invasive questionnaire features carried real but moderate information about CKD status in adults with long-standing diabetes in Bangladesh. The estimate is uncertain, the associations are cross-sectional, and two of them contradict clinical expectation, so the model is not ready for clinical use. At this sample size, single train/test splits and case-enriched samples can produce misleading accuracy and probabilities; repeated nested cross-validation with confidence intervals, together with external validation and recalibration, is needed before such models are used to prioritise testing.

## Data Availability

No new data were collected in this study. The analysed dataset, "A Dataset on Demographic and Lifestyle Factors for Prognosticating Chronic Kidney Disease Progression in Diabetic Patients" (version 2), is publicly available without restriction from Mendeley Data via https://doi.org/10.17632/hjkzgbxgv5.2 under a CC BY 4.0 licence. All analysis code, including the notebook that regenerates every number, table and figure in the paper, and the outputs of the reported run are available at Zenodo via https://doi.org/10.5281/zenodo.23758090 (version 1.0.0) and on GitHub at https://github.com/PanzerFaustAnan/diabetic-ckd-questionnaire-evaluation, under the MIT licence.

https://doi.org/10.5281/zenodo.23758090

https://doi.org/10.17632/hjkzgbxgv5.2

## Acknowledgments

The authors thank the patients who took part in the original data collection and the Department of Nephrology, BIRDEM General Hospital, for facilitating it.

## Supporting information

**S1 Checklist. TRIPOD+AI checklist.** Location of each reporting item in the manuscript.

**S1 Appendix. Supplementary tables and figures.** Table A: predictor definitions, coding and expected direction of association. Table B: variance inflation factors. Table C: observed CKD proportion by risk group. Table D: net benefit at selected thresholds. Table E: discrimination and calibration by subgroup. Table F: full specification of the final model. Table G: unpenalised multivariable logistic regression. Table H: sensitivity analyses. Table I: feature-group ablation. Table J: CKD proportion by diabetes management pattern. Table K: added value of the management pattern. Table L: within-patient change of time-varying features. Table M: landmark analyses. Table N: case-enriched sampling simulation. Table O: single-split versus repeated cross-validation simulation. Table P: public datasets. Fig A: subgroup performance. Fig B: management pattern. Fig C: within-patient change and landmark analyses. Fig D: learning curve. Fig E: single-split variability on public datasets.

